# Performance evaluation of Xpert HBV viral load (VL) assay: Point-of-care molecular test to strengthen and decentralize management of chronic hepatitis B (CHB) infection

**DOI:** 10.1101/2020.05.31.20104760

**Authors:** Khodare Arvind, Gupta Ekta, Nitiksha Rani, Singh Gaurav, Aggarwal Kavita, Sharma Manoj, SK Sarin

## Abstract

**Introduction:** Estimation of hepatitis B (HBV) viral load (VL) is critical in hepatitis-B cascade-of-care and currently there is no point of care (POC) molecular assay available for that. This study evaluated the performance of a new near point of care molecular assay Xpert HBV-VL assay against FDA approved Real time PCR assays.

**Materials & methods:** In this retrospective study 119 archived plasma samples from HBV infected patients, and 53 hepatitis B surface antigen (HBsAg) patients were simultaneously tested for HBV DNA quantification on 2 real time PCR conventional assays and Xpert assay. The routine method for reporting to patient was Abbott Real Time PCR.

**Results:** The range of HBV DNA load in samples was 1 to 8.76 log_10_IU/ml with a median load of 4.46 (IQR: 1-8.76) log_10_IU/ml as detected by routine assay (Abbott Real-Time HBV VL assay). Genotyping could be done in 95 (79.8%) samples and genotype D (83; 87.37%) was found commonest. The Xpert assay demonstrated good correlation with Abbott (R^2^= 0.944) and Roche (R^2^= 0.963). On comparison the mean difference (95% Confidence Interval) in average viral load was −0.018 log_10_ IU/ml and −0.043 log_10_ IU/ml when Xpert was compared with the Abbott and Roche assay, respectively. The overall sensitivity, specificity, negative predictive value and positive predictive value of the Xpert assay was found 97.5%, 100%, 94.65 & 100% respectively.

**Conclusion:** Xpert HBV-VL assay which has a potential for near point of care molecular testing has shown excellent performance and found to be a reliable method for HBV DNA quantification.

## Introduction

Infection with the hepatitis B virus (HBV) remains an important global public health problem with significant morbidity and mortality.[1] Globally the estimated prevalence of CHB infection is 3.5% with 257 million cases which accounts for 80% cases of hepatocellular carcinoma (HCC) along with chronic hepatitis C infection.[2] There is a large regional variation in hepatitis B seroprevalence between low (<2%) and high (>8%) endemicity levels. India falls in the intermediate endemicity zone (2%-8%) with an average prevalence of 4% (50 million cases) which is responsible for 43% of HCC cases.[3]

The primary diagnosis of hepatitis B is based on the detection of HBsAg but HBV DNA measurement is essential for the evaluation of patients with CHB for prognostication, to make decision to treat and subsequent monitoring of patients for the efficacy of antiviral treatment. For these indications serial monitoring of HBV-DNA levels is more important than any single arbitrary cut-off value. Interpretation of HBV-DNA levels is important in the context of other host factors including age, duration of infection, ALT elevation, and stage of disease when making treatment decisions.[4] The risk of disease progression is reduced when a sustained reduction of HBV DNA levels to undetectable is achieved, which in turn prevents the progression of fibrosis to cirrhosis, HCC and death.[5]

International practice guidelines for hepatitis B recommend using sensitive nucleic acid amplification test (NAAT) for quantifying HBV DNA in clinical practice.[1,4] There are several commercial NAAT assays, mostly real-time polymerase chain reaction (real-time PCR) are available for quantification of HBV DNA in clinical samples. [5–7] There are challenges in doing testing by conventional PCR methods Specialized infrastructure, trained manpower and longer turnaround time. [8,9]. A less complex, easy to use and inexpensive assay that has potential for point-of-care (POC) molecular testing are needed for more widespread availability of HBV management.

This study evaluated the performance of the new Xpert HBV VL assay (Cepheid, Inc. Sunnyvale, CA, USA) on the GeneXpert system (Cepheid) which is almost a POC or near POC test.

## Study design-

### Materials and methods

This was a retrospective study conducted in a tertiary care liver center of North India. Archived samples within 3 months of testing were retrieved from −80 °C and were simultaneously tested on all the three platforms from the same freeze-thaw cycle. Overall, 119 samples with confirmed HBV DNA positivity of CHB patients and 53 samples of HBsAg negative patients were included. Different technicians performed testing on the 3 platforms, and were blind to the results. The study was approved by Institutional Ethical Review. Samples from patients coinfected with HIV or HCV, children (<18 years old) and pregnant females were not included in the study and with insufficient available volume. Hepatitis B virus genotyping results were available for samples with DNA load >3log_10_ IU/ml and was done by Sanger sequencing method for the surface gene (S) of HBV genome.

### Xpert® HBV VL assay

This is an automated, real-time PCR assay for the quantification of HBV DNA performed on the GeneXpert System. The reported lower limit of quantification (LLOQ) is 1 log_10_ IU/ml and the linear range of quantification is 1-9 log_10_ IU/mL. The test requires 600 μL plasma or serum. All the steps like nucleic acid extraction, amplification, and detection of target done in a single cartridge and the result available in 59 minutes. The assay cartridge contains internal controls to ensure valid performance of the test and to quantify HBV DNA load by proprietary software. The GeneXpert System is available in a 1, 2, 4 or 16-module configuration. A four modules instrument was used for the present study.

### Abbott Real-Time HBV VL assay (Abbott, Wiesbaden, Germany)

This is an automated real-time PCR for the quantification of HBV DNA in human plasma or serum. The LLOQ is 1 log_10_ IU/mL and the linear range of quantification is 1-9 log_10_ IU/mL. The sample input volume is 500 μL of serum or plasma. Samples were tested on automated m2000sp-m2000rt Abbott real-time PCR.

### COBAS® AmpliPrep/COBAS® TaqMan® HBV Test, v2.0 (Roche Diagnostics, GmbH, Mannheim, Germany)

This is an automated HBV viral load quantitative assay. The LLOQ is 1.3 log_10_ IU/mL and the linear range of quantification is 1.3-8.23 log_10_ IU/ml. Sample input volume is 650 μL of serum or plasma. Test was performed on automated Roche COBAS Ampliprep/COBAS Taqman.

### HBV genotyping

HBV DNA extraction from plasma samples was done by a spin column-based method using the commercially available high pure viral nucleic acid kit (Roche Diagnostics, GmbH, Mannheim, Germany). The DNA extract was subjected to PCR using in-house designed primers for surface gene (S) genomic region (forward primer 5’-CATCAGGATTCCTAGGACCCCT-3’, reverse primer 5’-AGGACAAACGGGCAACATAC-3’) on Phusion® high fidelity DNA polymerase (Thermo Scientific Inc., Waltham, MA). Amplified products were purified by gel-excision using the QIAquick gel extraction kit (QIAGEN, GmbH, Mannheim, Germany) to remove unincorporated dNTPs and primers. This was followed by bidirectional sequencing using ABI Big Dye chemistry on the ABI 3500Dx series genetic analyzer (Life Technologies, Waltham, MA). Forward and reverse sequence reads were aligned and assembled using DNA Baser v3.5.1 software (Heracle BioSoft SRL, Romania). Genotype assignment was done by comparing the obtained sequences with the consensus sequences on the Basic Local Alignment Search Tool (BLAST) database of NCBI.

### Statistical analysis

The HBV DNA values were expressed in log_10_ format. Linear regression analysis was done and correlation coefficients were calculated using SPSS version 22. Agreement between the two assays was determined using Bland-Altman plots and estimated the overall bias. Sensitivity, specificity, positive predictive value (PPV), negative predictive value (NPV) was calculated by taking Abbott and Roche, both assay as reference.

### Results-

Of the 172 subjects included in the study, male preponderance was seen with M: F ratio of 3.3:1 and most were adults with a mean age of 43 (±14.7) years. The range of HBV DNA load in samples was 1 to 8.76 log_10_IU/ml with a median load of 4.46 (IQR: 3.12-6.39) log_10_IU/ml as detected by reference method (Abbott) which was used as a routine assay for reporting of results to the patients as per our institution’s practice. Genotype D was the commonest, seen in 83 (87.37%) of the cases.(Table 1) Median values of viral loads detected by all three assays are given in table 1.

**Table 1.** Baseline characteristics of the study population, n=172

| **Variable** | **Values** |
| --- | --- |
| **Age (Mean ± SD) years** | 43 ±14.7 |
| **Male gender, n (%)** | 132 (76.7%) |
| **Female gender, n (%)** | 40 (23.3%) |
| **Male: Female** | 3.3:1 |
| **ALT (median, IQR) IU/ml** | 45 (77-29) |
| **AST (median, IQR) IU/ml** | 41 (68-29) |
| **Genotype, n (%)** |  |
| **A** | 12 (12.63%) |
| **D** | 83 (87.37%) |
| **Median (IQR) HBV DNA (log<sub>10</sub> IU/ml)** |  |
| **Abbott** | 4.46 (6.39-3.12) |
| **Roche** | 4.47 (6.45-3.00) |
| **Xpert** | 4.35 (6.53-3.18) |
| **Xpert assay performance** |  |
| **Sensitivity** | 97.5% (95% CI; 92.8-99.5) |
| **Specificity** | 100% (95% CI; 93.3 -100) |
| **PPV** | 100% (95% CI; 96.9 -100) |
| **NPV** | 94.6% ( 95% CI; 85.1-98.9) |

**DNA: Deoxyribonucleic acid; IU: international unit, ALT: Alanine transaminase; AST: aspartate aminotransferase; SD: standard deviation; ml: millilitre. PPV & NPV: Positive & negative predictive value, CI: Confidence interval**

**Table 2.**
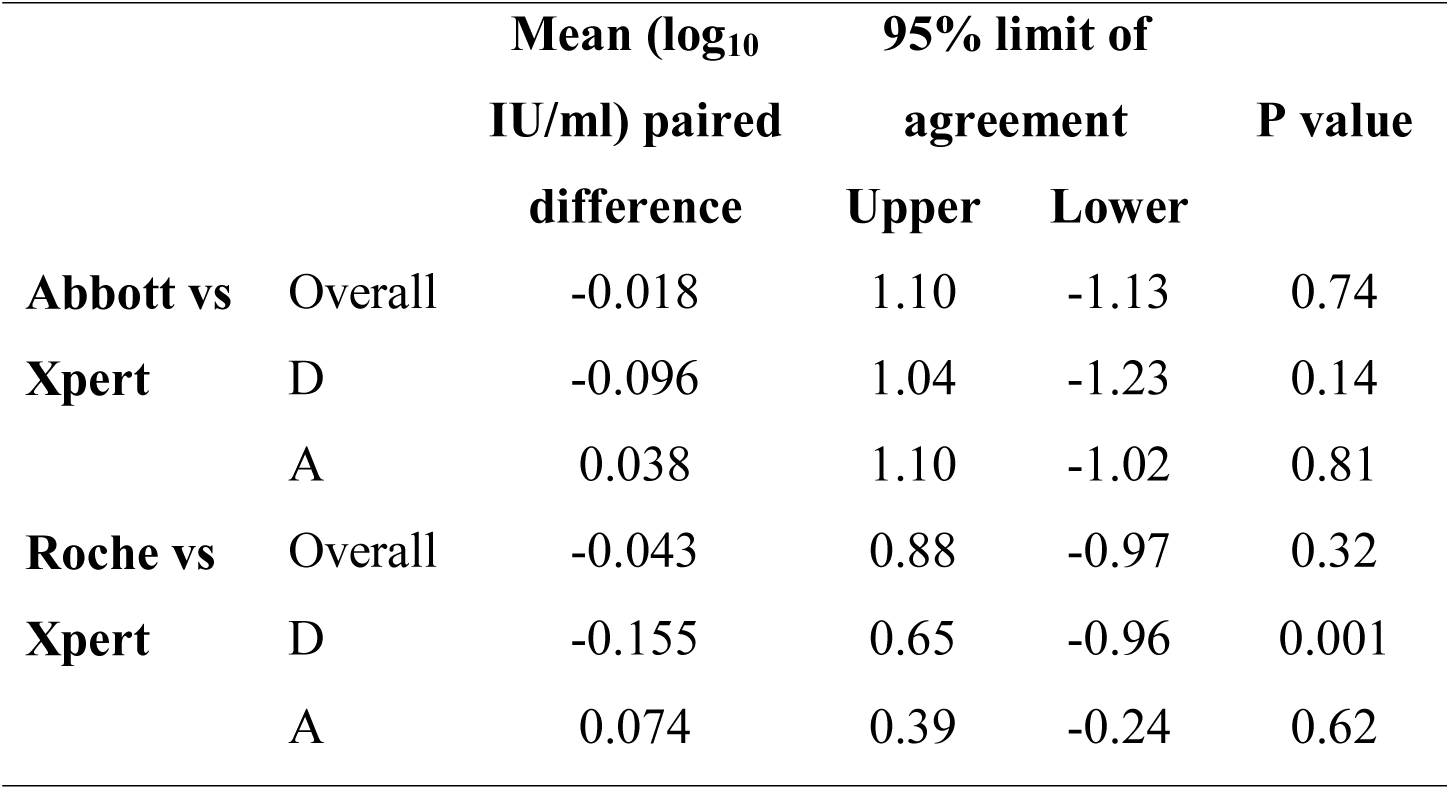
Level of agreement between the assays

**Table 3.** Repeatability of Xpert assay

| Number<br>of tests | High positive<br>standard<br>(4log<sub>10</sub>IU/ml) | Low positive<br>standard<br>(2log<sub>10</sub>IU/ml) |
| --- | --- | --- |
| **1** | 4.04 | 2.04 |
| **2** | 4.00 | 2.03 |
| **3** | 4.00 | 2.02 |
| **4** | 4.01 | 2.01 |
| **5** | 3.99 | 1.99 |
| **6** | 4.00 | 1.99 |
| **7** | 3.99 | 1.99 |
| **8** | 3.95 | 1.98 |
| **9** | 4.02 | 2.02 |
| **10** | 4.01 | 2.01 |
| **Mean** | 4.001 | 2.08 |
| **SD** | 0.023 | 0.020 |
| **CV%** | 0.56 | 0.99 |

### Comparison between Abbott and Xpert assay

On linear regression analysis of the quantifiable viral loads a very high positive correlation was seen (R^2^= 0.944) (Fig.1) and on Bland-Altman analysis 95% samples showed results within the limit of agreement of the assay ranging from −1.13 to 1.1 log_10_ IU/ml with a bias of −0.018 log_10_ IU/ml (Fig.2A).

**Figure 1.**
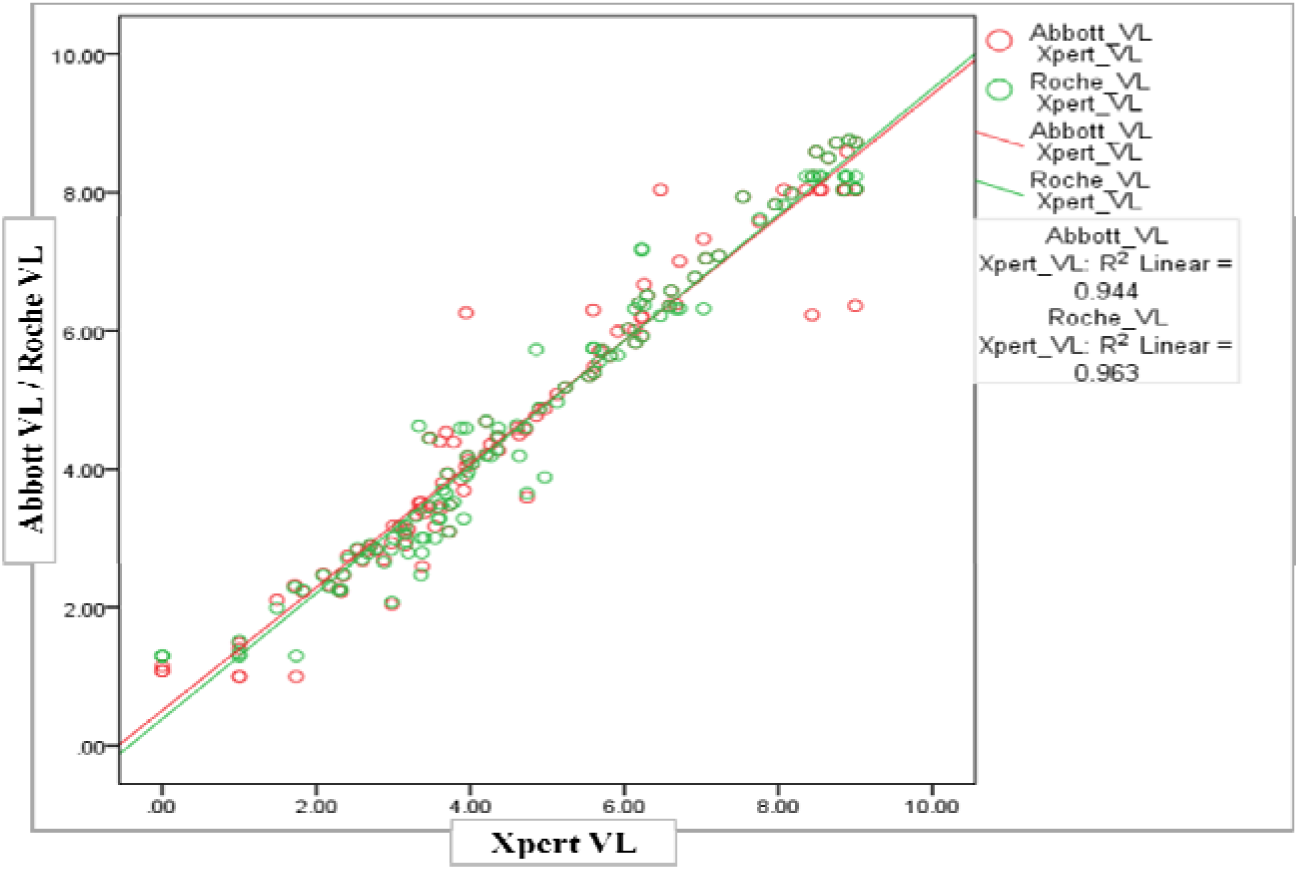
Linear regression analysis for correlation of Xpert assay with Abbott and Roche assay for quantification of HBV DNA.

**Figure 2.**
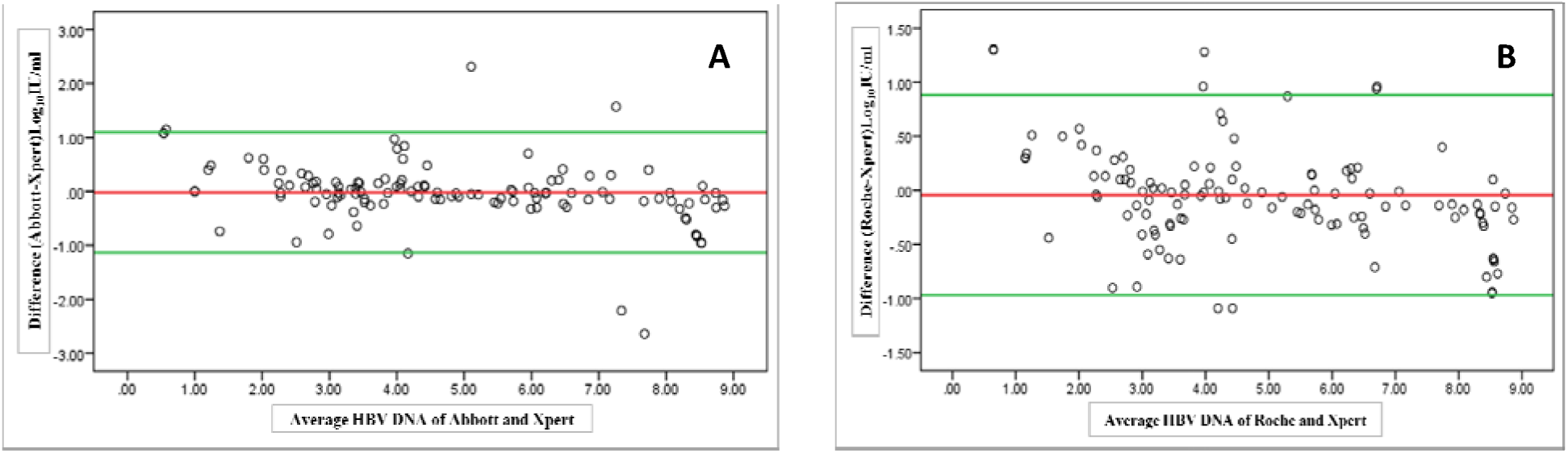
Level of agreement for quantification of HBV DNA between the assays by Bland-Altman plot; between Abbott and Xpert assay (A), between Roche and Xpert assay (B)

### Comparison between Abbott and Roche assay

On linear regression analysis of the quantifiable viral loads a very high positive correlation was seen (R^2^= 0.963) (Fig.1) and on Bland-Altman analysis 95% samples showed results within the limit of agreement of the assay ranging from −0.96 to 0.65 log_10_ IU/ml with bias of −0.043 log_10_ IU/ml (Fig.2B).

### Clinical performance of Xpert

Overall concordance was seen in 169 (98.25%) of 172 samples when Xpert assay was compared with Abbott and Roche assay. The sensitivity of Xpert assay was found 97.5% (95% CI; 92.8-99.5). All the 53 HBsAg negative samples tested negative by all three assays, so the specificity was found 100% (95% CI; 93.3 −100). There were 3 (2.52%) out of 119 samples had discrepant results and the viral load detected by Abbott assay in these three was 14, 12 and 12 IU/ml, but were not detected by Xpert. In these 3 samples, HBV DNA was also detected by Roche and it was less than the LLOQ i.e. 20 IU/ml. Due to insufficient sample volume the assays were not repeated in these 3 cases.

### Analytical performance of Xpert assay

To check analytical performance of Xpert assay 3^rd^ WHO International Standard for HBV (NIBSC code 10/264) was used.[10] The reconstituted standard was diluted in human plasma tested negative for all markers of HBV, hepatitis C virus (HCV) and HIV to make high positive (4 log_10_IU/ml) and low positive (2 log_10_ IU/ml) standard. For precision testing, high and low positive standards were tested on Xpert in duplicates on 5 consecutive days (to obtain 10 data points for each dilution) to evaluate the precision of the assay. The high positive standard was also used to make four serial 10-fold dilutions having a concentration of 4, 3, 2 and 1 log_10_IU/ml and tested to evaluate the linearity of Xpert assay for quantification.

The Xpert assay demonstrated good precision with a coefficient of variation (CV) for the ten HBV DNA load measurements which was 0.56% for the high positive standard and 0.99% for the low positive standard. (Table 3) On linear regression analysis of four serial 10 fold dilutions of WHO standard, an excellent correlation was seen between the measured HBV DNA concentrations and the expected concentrations (R^2^ = 0.998). (Figure 3) On analytical performance, the Xpert assay was found to be linear and reproducible.

**Figure 3.**
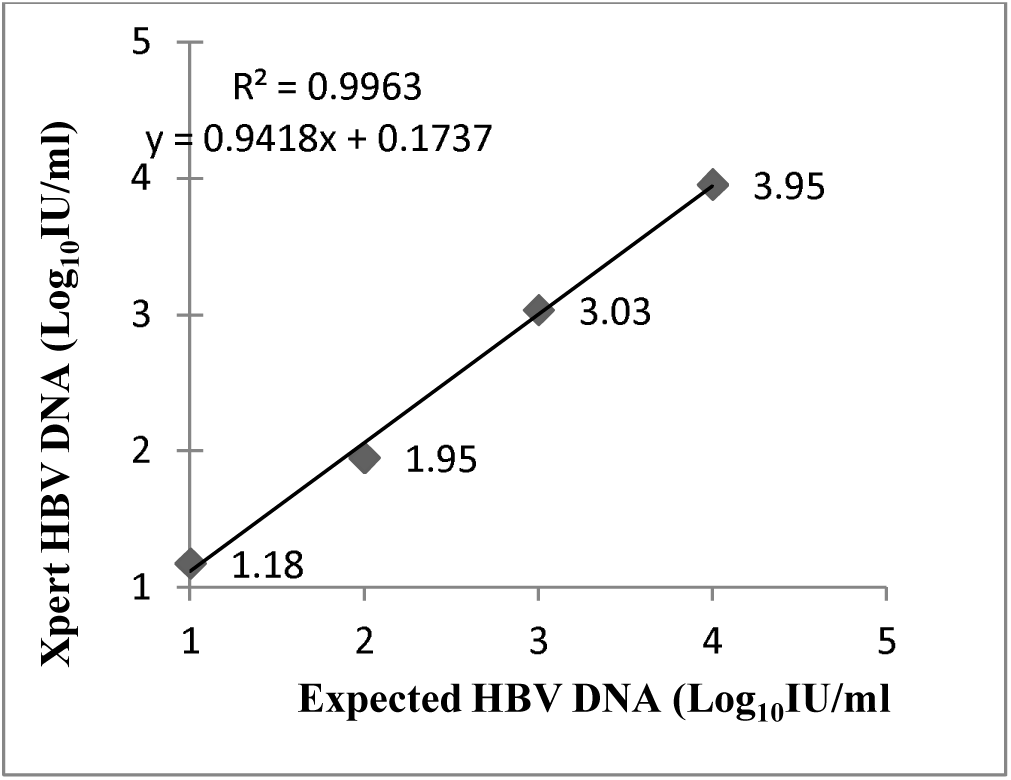
Xpert assay HBV DNA quantification linearity

**Figure 4.**
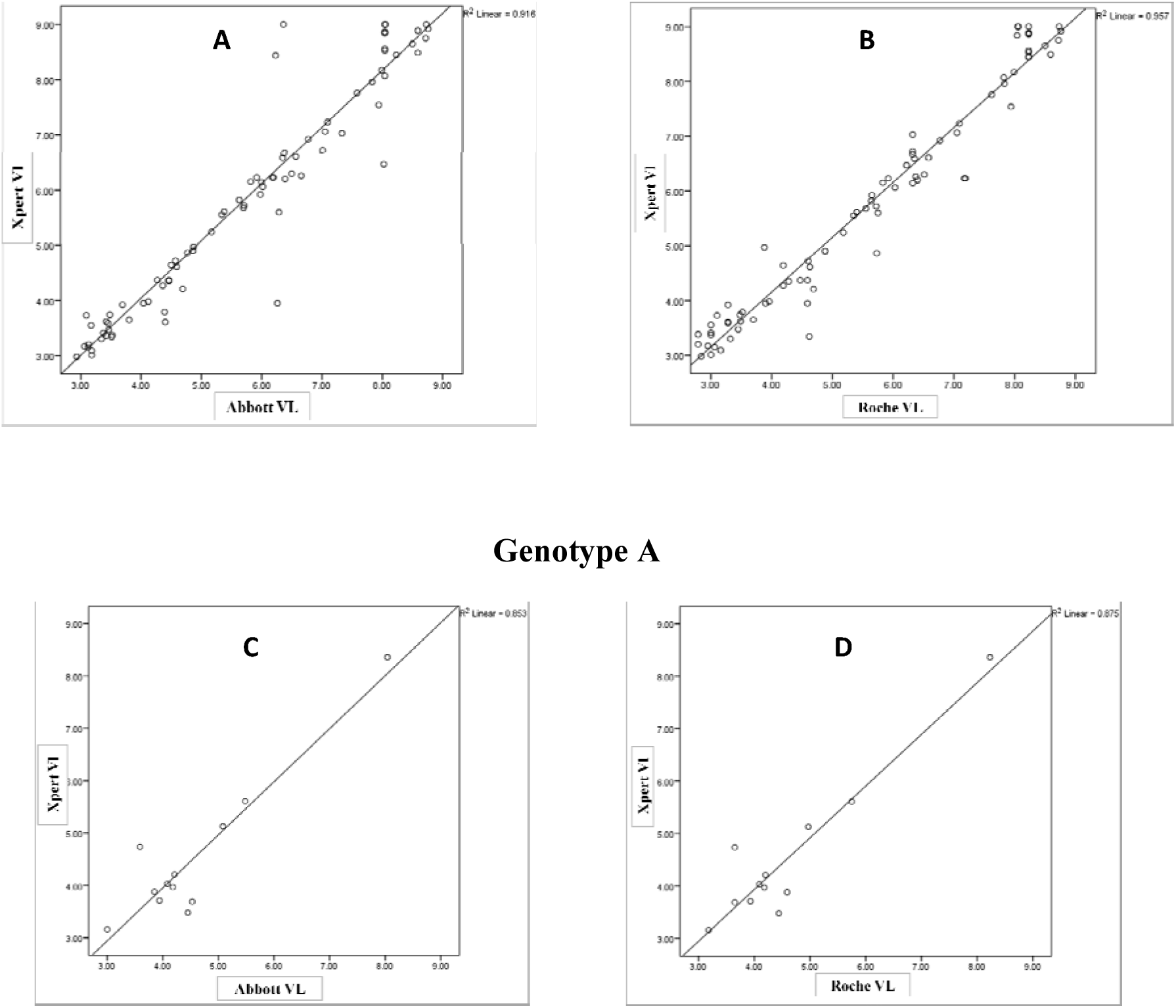
Correlation of measurements between the assays for HBV genotype D and A.

Figure 1 has shown the correlation of measurements between Abbott, Roche and Xpert assay. An excellent correlation between the Abbott and Xpert assay was observed (R2= 0.944, P < 0.001). An excellent correlation was also observed between the Roche and Xpert assay (R^2^ =0.963, P < 0.001).

Figure 2 A has shown the mean difference (Abbott – Xpert) of −0.018 log_10_IU/mL (limits of agreement: –1.13 to 1.1 log10IU/mL) and 2B has shown the mean difference (Roche – Xpert) of −0.043 log_10_IU/mL (limits of agreement: –0.97 to 0.88 log10IU/mL).

Figure 3 has shown the linearity of dynamic range of Xpert assay of quantification of HBV DNA (R^2^ =0.996)

Figure has shown the correlation of measurements between Abbott, Roche and Xpert assay for HBV genotype D and A. For genotype D a very good correlation between the Abbott and Xpert assay was observed (R2= 0.916, P < 0.001) (A). An excellent correlation was also observed between the Roche and Xpert assay (R^2^ =0.957, P < 0.001) (B).

For genotype A, a fairly strong correlation was found for Xpert assay when compare with Abbott (R2=0.853, P value <.001) (C) and Roche (R2=0.875, P value <.001) assay (D).

**Figure 5.**
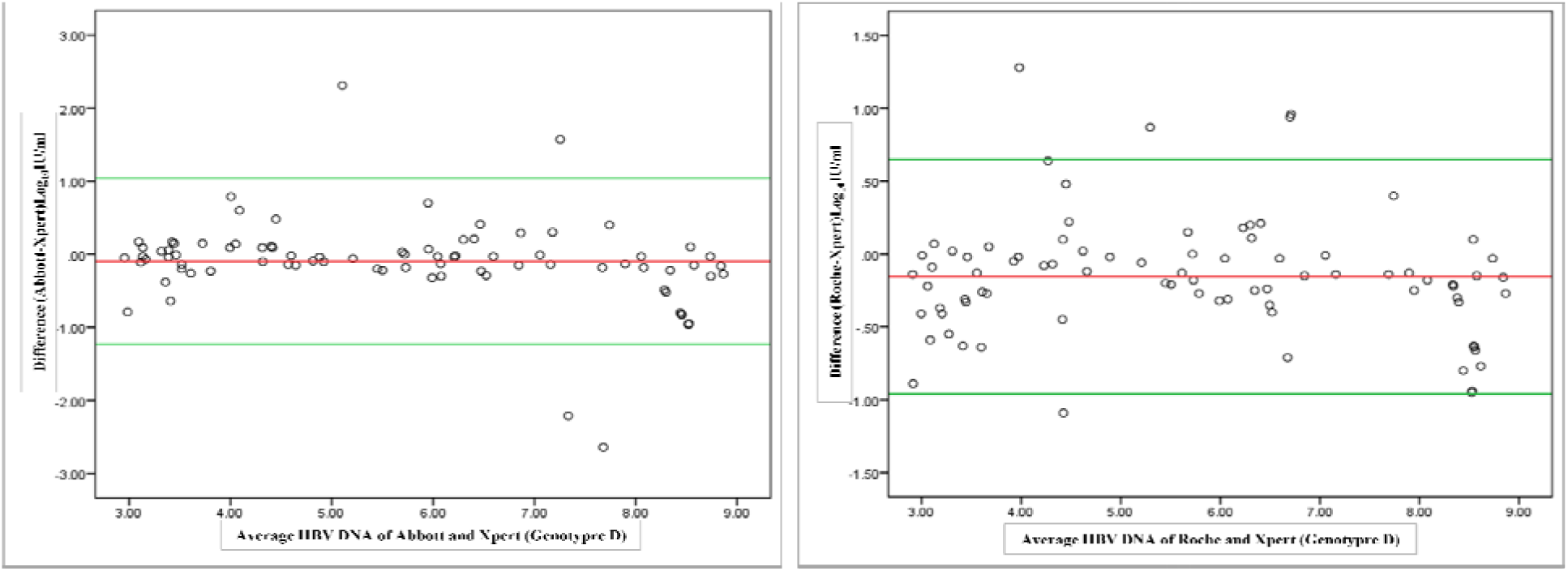
Level of agreement between the assays for samples with HBV genotype D and A.

For genotype D the mean difference was −0.096 (limits of agreement: –1.23 to 1.04) by Abbott and Xpert assay (A) and −0.155 log10IU/mL (limits of agreement: –0.96 to 0.65 log10IU/mL) by Roche and Xpert assay (B).

## Discussion-

Estimation of HBV-VL is critical in hepatitis-B cascade-of-care and at present, there is no POC molecular assay available for HBV-VL measurement. This study evaluated one such assay (Xpert assay) which has the potential to be used as a near POC molecular testing.

In this study, Xpert assay demonstrated very good specificity and sensitivity for the HBV DNA measurement as compared to FDA approved conventional assays.

There is only one published study that evaluated Xpert assay for HBV DNA testing against the Aptima assay (Hologic Inc. San Diego, CA, USA), a real-time transcription-mediated amplification (TMA), shown 100% sensitivity for samples with viral load > 10 IU/ml and 100% specificity. [9] Similarly we also did not found any false-positive results. A previous report found false-positive results with other real-time PCR assays but it was more frequent for assays that used manual extraction methods. [6] The simplicity and the single cartridge format of the Xpert assay curb the risk of sample cross-contamination as all the steps (extraction, amplification, and detection) take place within the same cartridge in a separate module on the GeneXpert instrument.

Available commercial real-time PCR assays are very sensitive and have much lower LOD, so very low values near to the LOD are often considered as false positive results.

In this study, we got 3 (2.52%) false-negative results with Xpert assay in samples with viral load near the LLOQ. In a similar study, authors found 20% (2 out of 10) false-negative results of the WHO standards with DNA load 5 IU/ml and they also demonstrated analytical sensitivity of 7.5 IU/ml for Xpert assay. [9] This discordance remains to be explained but in clinical practice, it rarely affects the management of CHB patients because HBV DNA is usually done to prognosticate CHB and initiate as well as to monitor antiviral treatment. As per the international guidelines the limit of HBV DNA used is quit high to characterize CHB and initiate antiviral treatment. [1,4] According to American Association for the study of liver diseases (AASLD) guideline (update 2018), in HBsAg and HBeAg positive patients treatment requires HBV DNA >20,000 IU/ml, elevated ALT (>2XULN) and/or moderate to severe inflammation or significant fibrosis detected either by liver biopsy or non-invasive methods of assessing fibrosis (elastography or FIB-4). However, HBsAg positive and HBeAg negative patients with HBV DNA level >2,000 IU/ml along with the other criteria (mentioned above) should be considered to start treatment.[4]

Assay reproducibility was excellent and the CV% was <1% for WHO standards with HBV DNA ≥2 log10IU/ml. One another study has also shown good reproducibility of Xpert assay.[9] The CV% for the low viral load was high and it was expected because for the low value any deviation from mean has shown more variation than the high values. When Xpert assays analyzed for different ranges of viral loads it demonstrated a very good correlation and level of agreement with reference assays irrespective of the viral genotype prevalent in India (genotype A and D), which was consistent with other studies.[7,9]

Currently, we have many molecular assays that have shown good clinical performance. Each has its advantages and drawbacks. As molecular assays like PCR requires specific infrastructure and these facilities are currently available at apex centers especially in resource-limited countries. So the diagnostic service delivery must be expanded to better manage the number of people living with HBV especially in peripheral locations. Currently available assays are although automated, uses several separate analytical steps, required several controls, expertise, specific infrastructure to perform efficiently and at least one full day of work to complete the procedure. Further, these molecular assays require batch testing to justify and to minimize the cost of the test which leads to the increase in turnaround time (TAT) which adversely increases the total visits of patients, dropouts, and also an economic burden. The Xpert assay has the potential to overcome all these drawbacks associated with available assays and may help to maximize the benefits of HBV-specific interventions by decreasing TAT, enhancing clinical service engagement, and patient retention.

Xpert HBV VL assay is a near point of care molecular test which is a demand for decentralization of diagnostic facilities. The relatively easy to use, require minimal training to perform and no other specific requirement for setup establishment make the Xpert HBV assay particularly well suited for use in resource-limited countries. Also very useful in developed countries because of its random excess, ability to avoiding batch testing coupled with its ability to deliver results fast. The maximum hepatitis B infected patients are living in low and middle-income countries[11] where clinical service coverage might be rudimentary. In these counties, GeneXpert platforms are already being used for diagnosis of tuberculosis, and it is also being used for quantification of HIV and HCV RNA [12–14], so it has the potential for an integrated testing approach and also it may increase access to HBV testing worldwide to incorporate people living with hepatitis B in the far-reaching areas.

The limitations of this study were that it was a single-center study and we did not have enough samples to retest discrepant results.

## Conclusion

Xpert HBV-VL assay has shown excellent performance and found to be reliable for HBV DNA quantification. It is a simplified, easy to use and has a potential for near POC molecular testing to expand service delivery for management of people living with HBV infection in centers with limited facilities and infrastructures.

## Data Availability

all the analyzed data has been included in the manuscript and the viral load of all individual tests has not mentioned.

## Acknowledgement

We acknowledge Cepheid (Inc. Sunnyvale, CA, USA) for kindly provided kits for testing.

